# Border control strategies for reducing importation risk of Clade Ib Mpox

**DOI:** 10.1101/2024.09.10.24313380

**Authors:** Shihui Jin, Tong Guan, Akira Endo, Gregory Gan, A. Janhavi, Gang Hu, Keisuke Ejima, Jue Tao Lim, Borame L Dickens

**Affiliations:** Saw Swee Hock School of Public Health, National University of Singapore and National University Health System, Singapore; Department of Infectious Disease Epidemiology and Dynamics, London School of Hygiene & Tropical Medicine, London, UK; School of Tropical Medicine and Global Health, Nagasaki University, Nagasaki, Japan; Centre for Mathematical Modelling of Infectious Diseases, London School of Hygiene & Tropical Medicine, London, UK; Lee Kong Chian School of Medicine, Nanyang Technological University

**Keywords:** Border measures, International travel, Mpox, PCR testing, Quarantine

## Abstract

**Background:** The Clade Ib monkeypox virus (MPXV), newly identified in the ongoing 2024 mpox outbreak, can be more transmissible through non-sexual routes compared to the previous Clade IIb strain. With imported cases sporadically reported globally, concerns have emerged about the potential of widespread transmission in the general community after importation events. Border control measures, such as screening and quarantining of arriving travellers, may help mitigate this risk and prevent localized outbreaks in the event of global spread.

**Methods:** We proposed nine border control strategies and evaluated their effectiveness in reducing importation risk using 10,000 microsimulations of individual infection profiles and PCR testing results under scenarios with varying disease prevalence levels (0.01%, 0.05%, and 0.1%) in the country of origin.

**Results:** The proposed border-control measures would reduce missed cases by 40.1% (39.1%–41.0%), 49.8% (48.8%–50.8%), and 58.1% (57.1%–59.0%) for predeparture, on-arrival, and both tests, respectively. Replacing the on-arrival test with a seven-day quarantine and post-quarantine testing would lower the count to 21.8% (20.9%–22.6%). Quarantine-only strategies showed a linear increase in effectiveness against duration, reaching a 90.4% (89.8%–91.0%) reduction with a 28-day quarantine. Disparities in distributions of missed case counts across strategies would become more pronounced at higher prevalence levels, with stringent approaches like quarantining followed by post-quarantine screening and 28-day quarantine keeping counts below two per 10,000 travellers, even at 0.1% prevalence.

**Conclusions:** When disease prevalence in the country of origin is low (0.01%), less restrictive approaches such as single on-arrival testing or a 14-day quarantine can maintain very low imported case counts of one or below. At higher prevalences, seven-day quarantining followed by post-quarantine testing, or 28-day quarantining is required to maintain similar effects. Decision makers will face balancing importation risk management and the negative impacts of such interventions to maintain safe international travel.

## 1. Introduction

Mpox is a viral zoonotic disease caused by the monkeypox virus (MPXV), typically characterized by symptoms such as skin rashes, fever, headache^1^. Clade Ib MPXV, the new strain in circulation in the ongoing 2024 mpox outbreak, is estimated to have emerged in late 2023 in the Democratic Republic of the Congo (DRC). This new clade has resulted in large number of infections in the DRC, and has spread to the neighbouring African countries, leading to over 100 confirmed cases in Burundi, Kenya, Rwanda, and Uganda as of 14 August 2024^2^. Cases of infections returning from Africa have also been identified and reported in countries outside the continent, such as Sweden and Thailand^3^.

While Clade IIb MPXV, the circulating strain in the 2022 mpox outbreak, was predominantly transmitted through sexual contact with minimal spread outside the community of men who have sex with men (MSM)^4,5^, the new Clade 1b has been found to transmit more easily than Clade IIb through non-sexual routes, such as skin-to-skin contact and contact with contaminated surfaces to some extent^6^.

Although transmissibility estimates have yet to be available, concerns have arisen on the potential for transmission in the general population globally^7,8^ due to its fast spread observed so far and the overall low immunity against mpox, particularly Clade Ib strain, in most countries outside the African continent.

At the frontline, border control measures can be employed to reduce importation rates and delay the epidemic peak^9^. Complete border lockdown introduces many social and economic challenges^10^, and considering the currently limited number of identified cases outside Africa, gives little imperative to do so. However, implementing less stringent containment approaches, such as screening and quarantining international arrivals from affected countries, can assist in the timely detection of imported cases and prevention of secondary infections, as well as provide treatment to infected travellers. Despite their successful application during the early phase of the COVID-19 pandemic^11^, few border control measures have been adopted in the previous mpox waves. Their application for mpox remains uncertain as mpox has different transmission timescales from COVID-19, including a substantially longer infectious period (14–28 days)^1,12^.

To assess strategies of cross-border control for mpox transmission, we propose a range of border control strategies and projected their effectiveness in reducing importation risks. We employed agent-based simulations to quantify the risk of importation per 10,000 travellers from countries with varying levels of disease prevalence. We explored the use of diverse containment measures, including quarantine and PCR testing before departure, upon arrival, and post-quarantine. We estimated and compared the number of missed and detected cases across a range of border control strategies at different disease prevalences (0.01%, 0.05%, 0.1%). Our findings aim to provide quantitative evidence for the designing of border control interventions for any country at risk of case importation globally, contributing as a united community to curb the global spread of mpox^13^ to support the focusing of interventions and treatment of communities currently affected.

## 2. Methods

An agent-based model for disease progression and testing was employed to generate individual infection profiles, testing procedures and outcomes under various border control measures. Details of the models and the proposed containment strategies are described in sections below.

### 2.1 Model of disease progression

The infection process was simulated independently for each infected traveller, who was assumed to contract the virus at some point within 30 days prior to departure. We used the time from departure for all the travellers as time scale,. For an infected traveller, his or her infection time point was denoted as, followed by an incubation period and an infectious period. We assumed no pre-symptomatic infection. Both and were modelled using log normal distributions to account for their heavy-tailed nature, with parameters derived from statistics disclosed by WHO^1^ (Figure 1, Table 1).

**Table 1.** Parameters used within the model.

| Variables | Value, range or distribution | Definition |
| --- | --- | --- |
| $\tau_i$ | Uniform distribution ranging from -30 to 0 | Day of infection of individual $i$ before departure |
| $L$ | Lognormal distribution with mean 8.72 and standard deviation (SD) 4.44 | Incubation period duration of individual $i$; derived from WHO<sup>1</sup> |
| $\varphi_i$ | Lognormal distribution with mean 20.11 and standard deviation (SD) 3.58 | Infectious period duration of individual $i$; derived from WHO<sup>1</sup> |
| $\sigma_{skin}(t)$ | Function of PCR testing sensitivity against time from symptom onset, determined from logistic regression | Post infection time-dependent sensitivity of PCR test using samples of skin lesions; derived from Yang *et al.*<sup>14</sup> |
| $\sigma_{oral}(t)$ | Function of PCR testing sensitivity against time from symptom onset, determined from logistic regression | Post infection time-dependent sensitivity of PCR test using oropharyngeal samples; derived from Yang *et al.*<sup>14</sup> |
| $X_i$ | Binary, i.e., 0 or 1 | Indicator for positive PCR test result |
| $S_i$ | Binary, i.e., 0 or 1 | Indicator for developing skin rashes |

**Figure 1.**
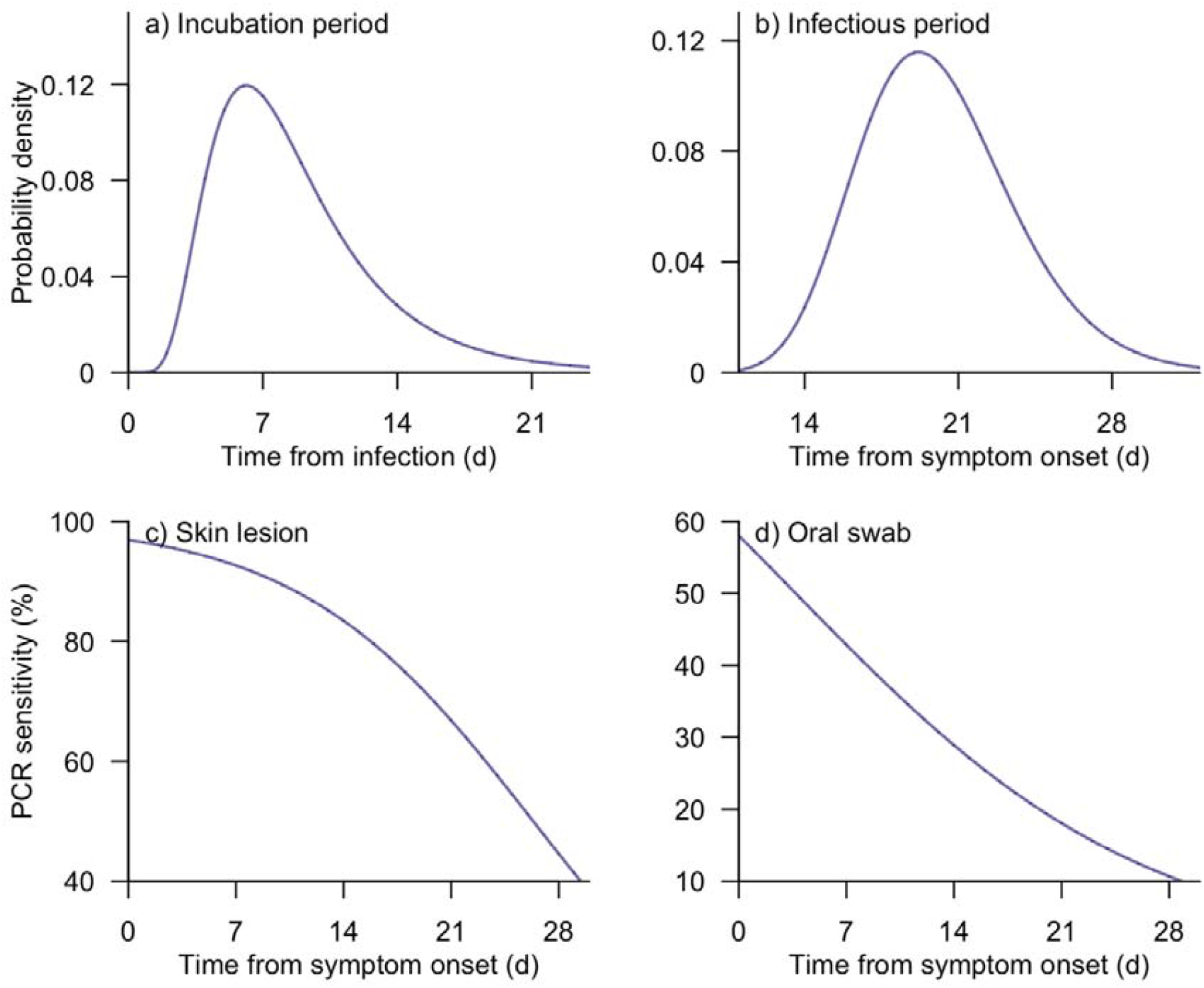
Distribution of the parameters for the model of disease progressionHH. Probability density for incubation period (a) and time from symptom onset to loss of infectiousness (b)^1^. Sensitivity of PCR test using samples of skin lesions (c) and oropharyngeal samples (d) over time^14^. Note that the time points with relatively low probability density (a–b) or sensitivity (c––d) may not be fully displayed.H

### 2.2. Model of PCR testing

The PCR testing methods considered in the simulations include sampling from skin lesions and oropharyngeal sites. Test sensitivity over time for the two PCR types, denoted as *σ_skln_* and *σ_oral_*, was inferred utilizing a logistic regression model and the test outcome data from Yang et al. (Figure 1, Table 1), in which PCR positivity was defined as viral load exceeding 4.77 log_10_ copies per mL^15^. Given the higher PCR testing accuracy when samples from skin lesions were used, we assumed that infected individuals who developed skin rashes (S_i_=1), accounting for 60% of all infections, would be tested with skin lesion swabs, while others (S_i_ = 0)would provide oropharyngeal samples for PCR testing. The probabilities of correctly identifying an infection in the two groups at testing time *t*_*i*_ are assumed to be

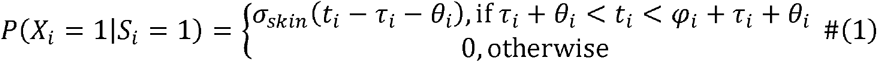

and

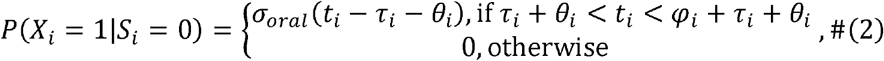

respectively. In these, we assumed that pre-symptomatic infections would evade PCR detection but tested the assumption in a sensitivity analysis (Figure S1). A turnaround of three days was assumed to account for laboratory processing and administrative procedures^16^. False-positive results were not considered in our simulations.

### 2.3 Proposed strategies

The border control measures considered in this study include pre-departure testing, and on-arrival testing, post-arrival quarantine, and post-quarantine testing. Pre-departure tests would be performed three days prior to departure with travellers not subject to any activity restrictions during this period. Only those with a negative test result would be permitted entry into the destination country. On-arrival testing would be conducted upon arrival and incoming travellers would be quarantined in designated facilities for three days whilst waiting for their test results. Individuals with a positive result would be transferred to local healthcare facilities for isolation and treatment, while others would be released into the community. Post-arrival quarantine would last between seven and 28 days, during which no test would occur. Post-quarantine testing would take place at the end of the quarantine period. Travellers testing negative would be released following an additional three-day quarantine due to test result delays, while those testing positive would also be isolated and treated in healthcare facilities until recovery. In addition, complete adherence was assumed during quarantine, isolation, or treatment process with no leakage expected.

The baseline strategy was set to be the one without screening or quarantine (i.e., Strategy 1), where all international arrivals were granted entry and free movement upon arrival. Eight other strategies, featuring different combinations of the four border control measures, were also explored in this study. These nine strategies are visualized in Figure 2 and listed as below:

**Figure 2.**
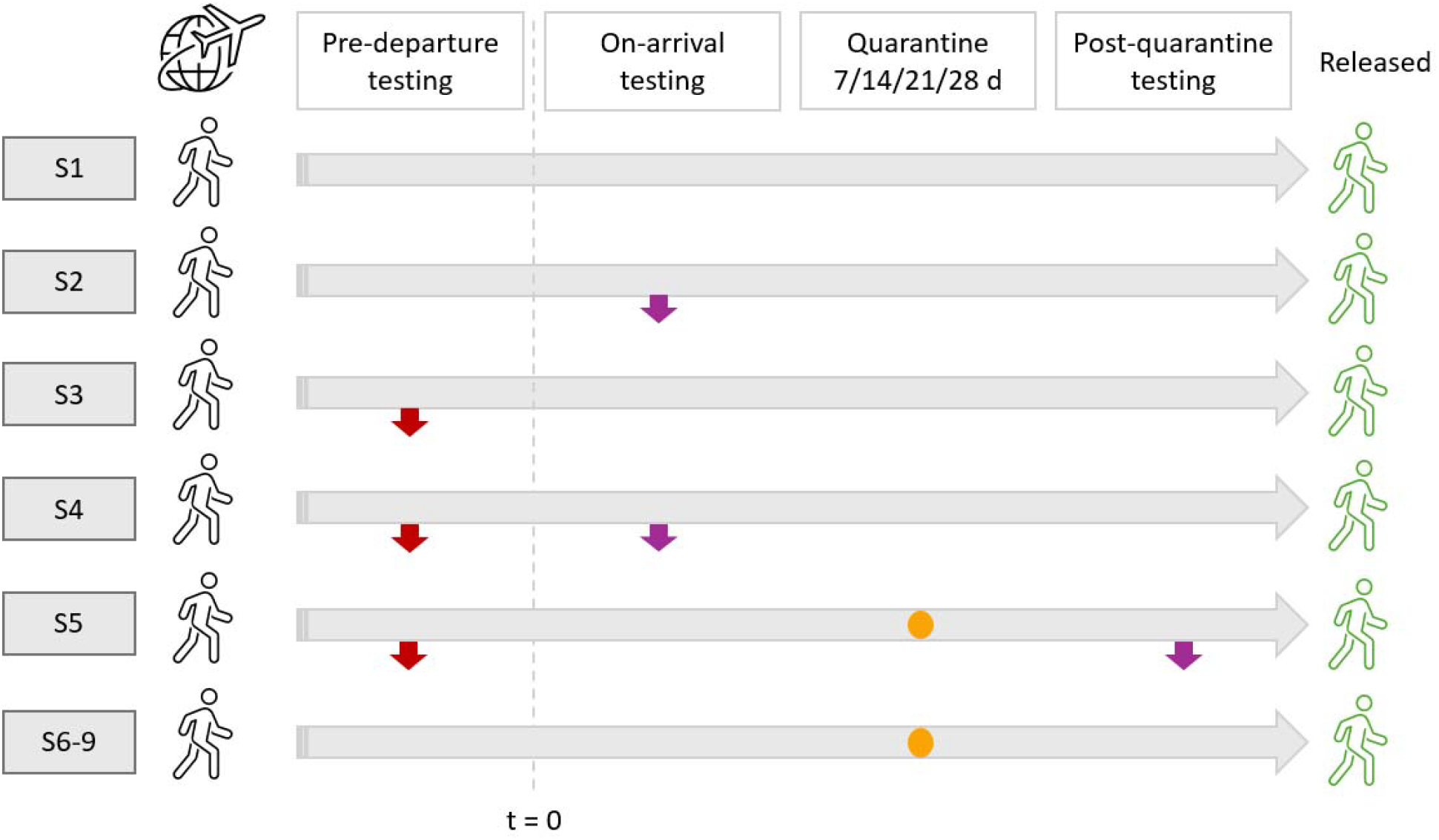
Descriptions of Strategies 1–9 and outcomes. Red arrows in Strategy 3–5 represent infected individuals detected by pre-departure PCR tests and thereby not permitted entry. Purple arrows in Strategy 2, 4, and 5 represent infected individuals detected by PCR tests or those who lost infectiousness while awaiting testing results. These individuals would not be able to infect others upon release into the community. Orange dots in Strategy 5–9 denote undiagnosed infections who lost infectiousness during quarantine. Strategies 6–9 are presented together in the last row as they differ only in quarantine duration. The time scale () is the time from departure for all the travellers.

S1) No screening or quarantine,

S2) On-arrival testing,

S3) Pre-departure testing,

S4) Pre-departure testing + on-arrival testing,

S5) Pre-departure testing + post-arrival quarantine (7 days) + post-quarantine testing,

S6) Post-arrival quarantine (7 days),

S7) Post-arrival quarantine (14 days),

S8) Post-arrival quarantine (21 days), and

S9) Post-arrival quarantine (28 days).

We quantified the effectiveness of individual strategies in reducing importation risks through proportion of missed cases, defined as individuals who were infectious or still in their incubation period by the time they were able to interact with the local community. These summary statistics were derived from the outputs of 10,000 microsimulations per strategy, each with 10,000 infectious or exposed arrivals. We further explored three disease prevalence levels, including 0.01%, 0.05%, and 0.1%, among travellers from the source region. For each border control strategy and disease prevalence level, we summarized the number of missed cases per 10,000 travellers, based on 10,000 simulations of 100,000 arrivals per scenario. All the analyses and result visualizations were performed with R software^17^.

## 3. Result

### 3.1 Overall effectiveness of proposed border control strategies

The simulation results showed that pre-departure PCR testing (S3) and on-arrival PCR testing (S2) could detect 40.1% (95% Confidence Interval [CI]: 39.1%–41.0%) and 46.2% (95% CI: 45.2%–47.2%) of all imported cases, respectively. In addition, 3.6% (95% CI: 3.3%–4.0%) of the infected travellers would lose infectiousness during the three-day quarantine while awaiting the on-arrival PCR test results. Combining both pre-departure and on-arrival testing (S4) would reduce the proportion of missed cases by an additional 8.3 (95% CI: 7.7–8.8) or 18.0 (95% CI: 17.3–18.8) percentage points (pp) compared to scenarios with only on-arrival or pre-departure testing, respectively. A further reduction of 20.2 (95% CI: 19.2–21.1) pp in the proportion of missed cases was expected when the on-arrival testing in S4 was replaced with a seven-day post-arrival quarantine and post-quarantine PCR testing (S5). In this scenario, while 28.5% (95% CI: 27.6%–29.4%) of the infected travellers were captured through post-quarantine testing in addition to the 40.1% (95% CI: 39.1%–41.0%) captured from pre-departure testing, another 9.7% (95% CI: 9.1%– 10.3%) turn non-infective during the 10-day quarantine, which included a three-day PCR test turnaround time. When quarantine was implemented as the exclusive border control measure, 20.7% (95% CI: 19.9%–21.5%), 24.8% (95% CI: 23.9%– 25.7%), 25.3% (95% CI: 24.5%–26.2%), and 19.5% (95% CI: 18.7%–20.3%) of infected travellers would lose infectiousness with each additional week of quarantine (Figure 3).

**Figure 3.**
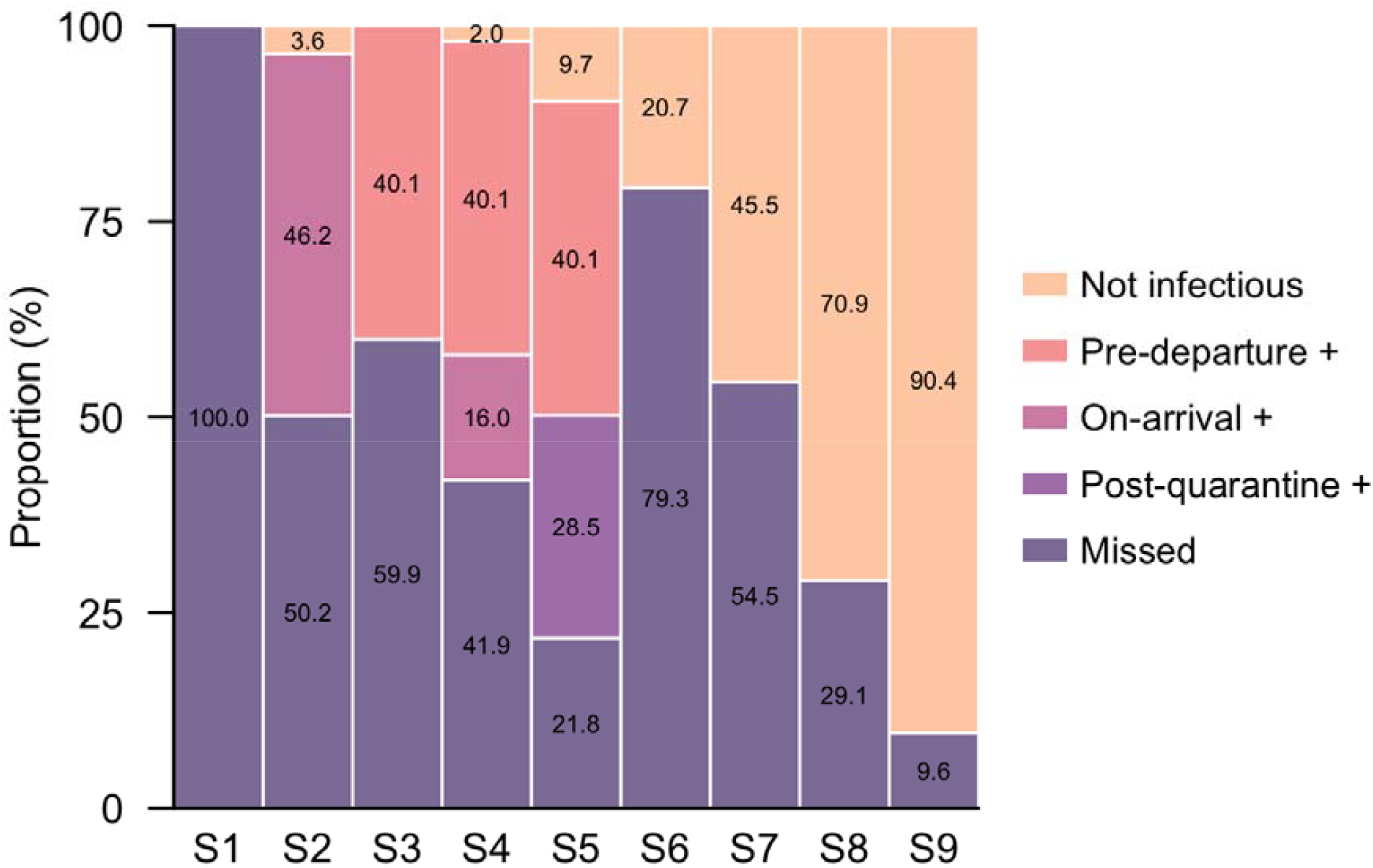
Proportion of cases missed, detected by PCR tests, and losing infectiousness during quarantine among infected travellers across Strategy 1–9. The five sections in the legend bar represent cases who lost infectiousness during quarantine or when awaiting PCR results (‘Not infectious’), cases testing positive before departure (‘Pre-departure +’), upon arrival (‘On-arrival +’), and post-quarantine (‘Post-quarantine +’), as well as cases missed and leaked into the (‘Missed’), respectively.

### 3.2 Number of missed cases across scenarios with varying disease prevalence levels

When the disease prevalence in the country of origin was low (0.01%), 53.9% of the 10,000 simulations indicated at least one missed case per 10,000 travellers in the baseline scenario without interventions. The corresponding risk was reduced to 2.9%, 8.3%, and 1.0% for the scenarios implementing solely on-arrival testing (S2), pre-departure testing (S3), and both (S4), respectively, while the substitution of the on-arrival testing in S4 with a seven-day quarantine and post-quarantine testing (S5) further lowered the risk to 0.02%. By comparison, in the quarantine-only scenarios, the risk was 26.8%, 4.8%, 0.11%, and 0 when the quarantine duration was seven, 14, 21, and 28 days, respectively (Figure 4).

**Figure 4.**
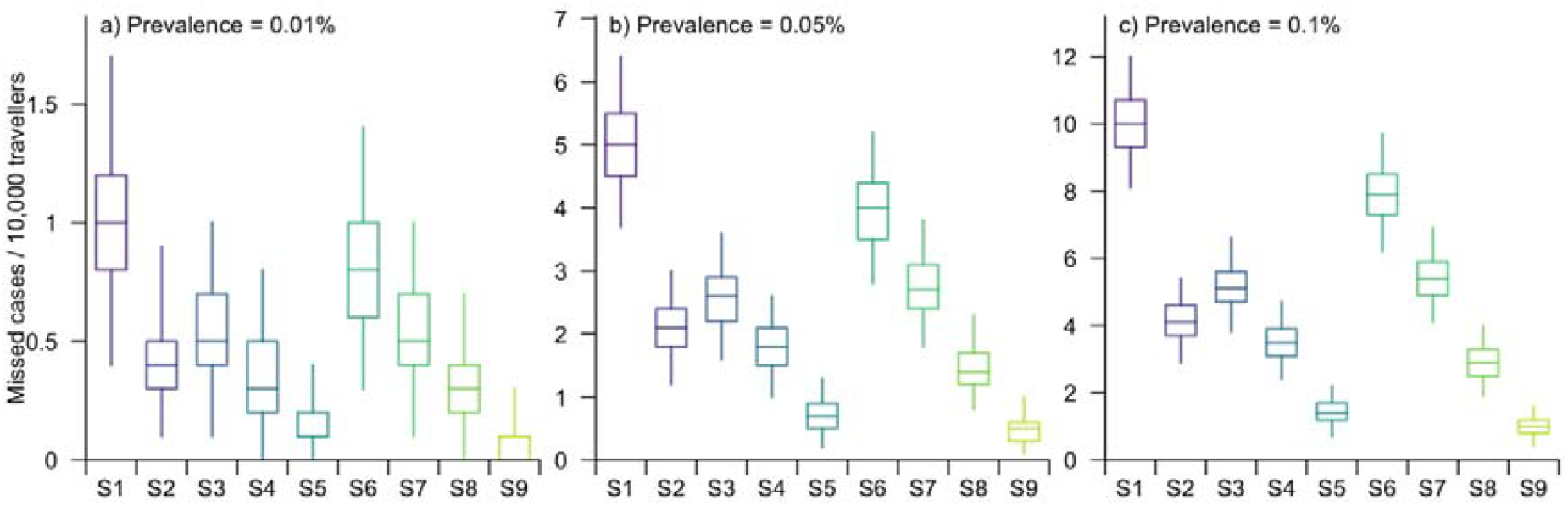
Number of missed cases per 10,000 travellers. Summary statistics include medians, interquartile ranges (IQRs), and 95% confidence intervals (CIs) for Strategy 1–9 when the disease prevalence was 0.01% (a), 0.05% (b), or 0.1% (c) in the country of origin.

As the disease prevalence increased to 0.05%, the number of missed cases per 10,000 travels in the baseline scenario was projected to be 5 (Interquartile Range [IQR]: 4.5 – 5.5). Performing PCR testing before departure, upon arrival, or at both time points would reduce the case count to 2.5, 3.0 and 2.1, with IQRs of 2.2 – 2.8, 2.6 – 3.4, and 1.8 – 2.4, respectively. Adopting the strategy with a seven-day quarantine followed by post-quarantine testing would further decrease the number of missed cases to 1.1 (IQR: 0.9 – 1.3). In comparison, the 7, 14, 21 and 28-day quarantine without testing would lower the number to 3.9 (IQR: 3.5 – 4.4), 2.7 (IQR: 2.4 – 3.1), 1.4 (IQR: 1.2 – 1.7), and 0.5 (IQR: 0.3 – 0.6), respectively. Similar trends were observed when the disease prevalence in the country of origin reached the hypothetical peak of 0.1%. Strategies which performed well in the lower-prevalence scenarios continued to substantially reduce the number of missed cases. Particularly, the count of missed cases was 2.2 (IQR: 1.9 – 2.5) for the strategy requiring pre-departure PCR test and a seven-day quarantine, 5.4 (IQR: 5.0 – 5.9) for 14 days’ quarantine, and 0.9 (IQR: 0.7 – 1.2) for 28 days’ quarantine (Figure 4).

## Discussion

Our results demonstrated significant reductions in the number of missed cases across all eight border containment strategies compared to the baseline, supporting the implementation of screening and/or quarantine measures at the border to mitigate the risk of further mpox transmission in the local community.

The comparison between timing of PCR testing showed higher effectiveness of on-arrival tests than pre-departure in reducing missed case counts. This difference could be partially attributed to the fact that some infectious arrivals lost infectiousness during the three days’ quarantine while waiting for test results. These individuals, who were in the late stages of infection, would likely not have been captured by pre-departure screening (Figure 1). More importantly, the delay in testing allowed the detection of some arrivals who were yet to be symptomatic three days before departure to become infectious by the time they arrived at the border. Postponing testing until seven days post-arrival would further inflate the sizes of the two groups, thereby lowering the number of missed cases. Meanwhile, the decrease in the number of missed cases for the strategies involving two rounds of PCR testing could also be explained by the reduced likelihood of false negatives, as dual tests would increase chances of detecting infections missed by a single test^18^.

The effectiveness of quarantine was projected to increase linearly with duration. While a seven-day quarantine provided only modest benefits in reducing importation risk, extending the period to 28 days lowered the risk by over 90%. This is primarily due to the long incubation and infectious periods of mpox, often requiring over three weeks from exposure to full recovery^19,20^. Although the quarantine-only strategies have various advantages in addition to containing importation, such as little demand and costs for PCR testing, as well as the possibility of utilizing existing infrastructure established during the COVID-19 pandemic to accommodate the incoming travellers^21,22^, its adverse impact on individuals’ mental health is of concern^23,24^. In addition, adopting long quarantining periods could economically harm many countries that rely on tourism as a major source of national revenue^24,25^. These downsides collectively can make long-duration quarantine unsustainable in the long run.

Through balancing the minimisation of importation size of infected travellers and alleviating the negative secondary effects caused by border restrictions, differentiated border management policies based on prevalence levels in countries of origin can be adopted^26^. The marginal distinctions in effectiveness between various testing schemes or quarantining, such as single on-arrival testing or 14-day quarantine for arrivals from countries with low disease prevalence, also provides options depending on available testing and isolation facility infrastructure. In contrast, for travellers from countries with higher prevalences (e.g., >0.05%), more stringent approaches, such as a seven-day quarantine followed by post-quarantine screening or a quarantine with a minimum length of 21 days, may need to be employed to minimise the disease importation risk. The continuation of increased efforts and support to estimate outbreak sizes and prevalence in affected countries as part of a robust global surveillance system for real-time monitoring of transmission could facilitate the timely adjustment of countries at risk of exporting infections and their level of risk^27^.

It should be noted that directly comparing the effectiveness of PCR testing and quarantine requires caution owing to the large uncertainties surrounding the testing sensitivity. At this time, uncertainties lie on the proportion of infections who develop skin rashes, which render the use of skin lesion swabs that substantially reduce the probability of false negatives^28^. In the main analysis, we assumed this proportion to be 60% but assessed in a subsequent sensitivity analysis how variations in this parameter could influence the effectiveness of border control strategies (Figure S2– S4). When fewer cases presented with skin rashes, PCR testing did not outperform long-term quarantine. Furthermore, the effectiveness of PCR testing could be undermined by fluctuations in accuracy across different testing kits or protocols applied in different locations^14^. Such potential inconsistencies, together with the resource demands of PCR testing, particularly for repeated tests, might make the quarantine-only strategies a more feasible and reliable option in controlling border risks.

There are a few other limitations in our study worth noting. These include the paucity of key parameters necessary for our simulation model, particularly for the latest Clade Ib MPXV. We relied on existing statistics for Clade IIb MPXV and general MPXV to derive parameters related to mpox infection progression, including distributions for incubation and infectious periods, as well as PCR testing sensitivity, assuming similar patterns would apply to the new Clade Ib strain. The insufficient data available on disease prevalence in each country and international travel volumes posed great challenges in accurately quantifying the burden of these strategies on quarantine facilities and number of PCR tests required at different time points. In addition, we assumed no infections occurred during the travelling period.

We also presumed complete adherence to quarantine protocols and no-cross infection among quarantined individuals. Although there has been limited evidence that mpox spreads through air, transmission via contaminated surfaces, apart from close contacts, remains possible^29^, making it difficult to rule out this risk.

Despite these limitations, our study provides an evaluation of the effectiveness of nine border control strategies on managing the importation risk of Clade Ib MPXV across different levels of disease prevalence in the country of origin. The projected outcomes of the proposed methods, including quarantine and screening at the border, demonstrate the potential of maintaining the risk at a manageable level. The varying intervention effects based on the disease prevalence in the country of origin underscore the imperative of implementing tailored border policies which account for mpox outbreak scales. Such differentiated strategies would help mitigate the negative impacts of interventions on individual well-being, social resources, and the economy, while effectively preventing a potential importation-driven outbreak.

## Data Availability

Data available upon request.

## Data Availability

Data available upon request.

## Author contributions

SJ, TG, and BLD conceived and designed the study. SJ and TG implemented the statistical analysis and created the figures and tables. SJ wrote the original draft of the manuscript. TG, AE, GG, AJ, GH, KE, JTL, BLD reviewed and edited the manuscript.

