## Supplementary for "Border control strategies for reducing importation risk of Clade Ib Mpox"

**Supplementary information**

### Impact of viral shedding before symptom onset on testing sensitivity

Given the potential for viral shedding before symptomatic onset^1^, we performed a sensitivity analysis to evaluate its impact on the effectiveness of the border control strategies. Specifically, we randomly selected ~10% of the infections to become infectious up to two days before symptoms appeared. As they did not develop any symptoms in this period, PCR tests was administered using oral swabs. Their testing results were simulated using the extrapolated sensitivity rates. We then calculated the proportions of missed cases, PCR detected cases, and those losing infectiousness during quarantine or while awaiting PCR results, among all infected travellers. The results showed only marginal differences compared to the main analysis, which assumed no pre-symptomatic viral shedding detectable by PCR (Figure S1).

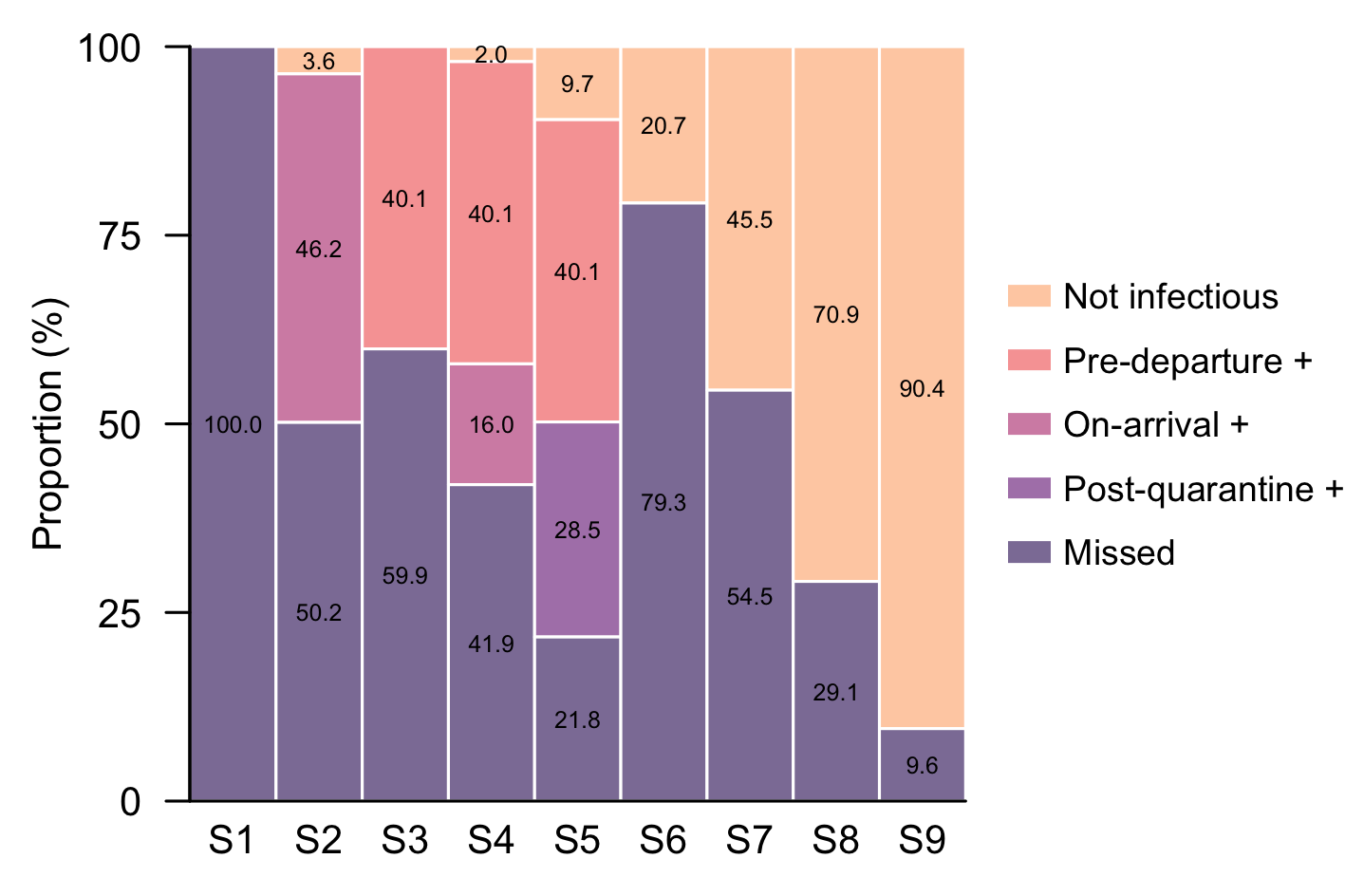

**Figure S1.** Proportion of cases missed, detected by PCR tests, and losing infectiousness during quarantine among infected travellers across Strategy 1–9, when viral shedding before symptom onset was considered. The five sections in the legend bar represent cases who lost infectiousness during quarantine or when awaiting PCR results (‘Not infectious’), cases testing positive before departure (‘Pre-departure +’), upon arrival (‘On-arrival +’), and post-quarantine (‘Post-quarantine +’), as well as cases missed and leaked into the (‘Missed’), respectively.

### Impact of PCR swab types on testing sensitivity

Since PCR testing sensitivity depends on the type of swabs used^2^, we assumed in our simulations that all infections who developed skin rashes would undergo PCR testing using samples from skin lesions, while others would be tested using oral swabs. However, the exact proportions of these two subpopulations remain unknown. In the main analysis, we presumed the proportion of those with skin rashes to be 60%, but here we assessed the effectiveness of various intervention strategies in reducing missed case proportions under alternative assumptions. We considered three candidate values 10%, 30%, and 90%, and the results are shown in Figure S1–S3. A reduction in the proportion of missed cases was observed with the increase in the proportion of infections with skin rashes, but the relative effectiveness of strategies with different testing schemes remained consistent.

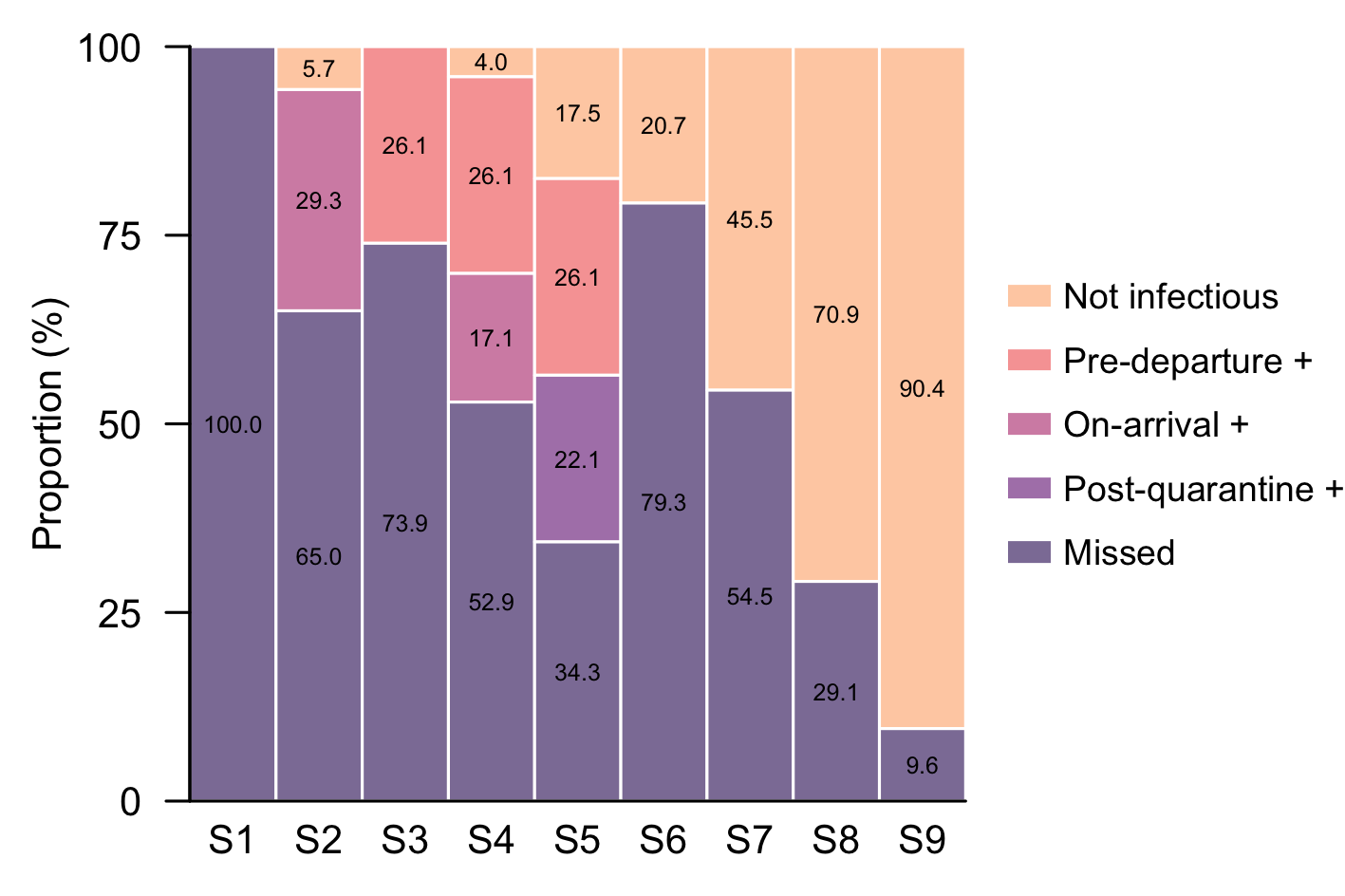

**Figure S2.** Proportion of cases missed, detected by PCR tests, and losing infectiousness during quarantine among infected travellers across Strategy 1–9, when the proportion of infections with skin rashes was 10%. The five sections in the legend bar represent cases who lost infectiousness during quarantine or when awaiting PCR results (‘Not infectious’), cases testing positive before departure (‘Pre-departure +’), upon arrival (‘On-arrival +’), and post-quarantine (‘Post-quarantine +’), as well as cases missed and leaked into the (‘Missed’), respectively.

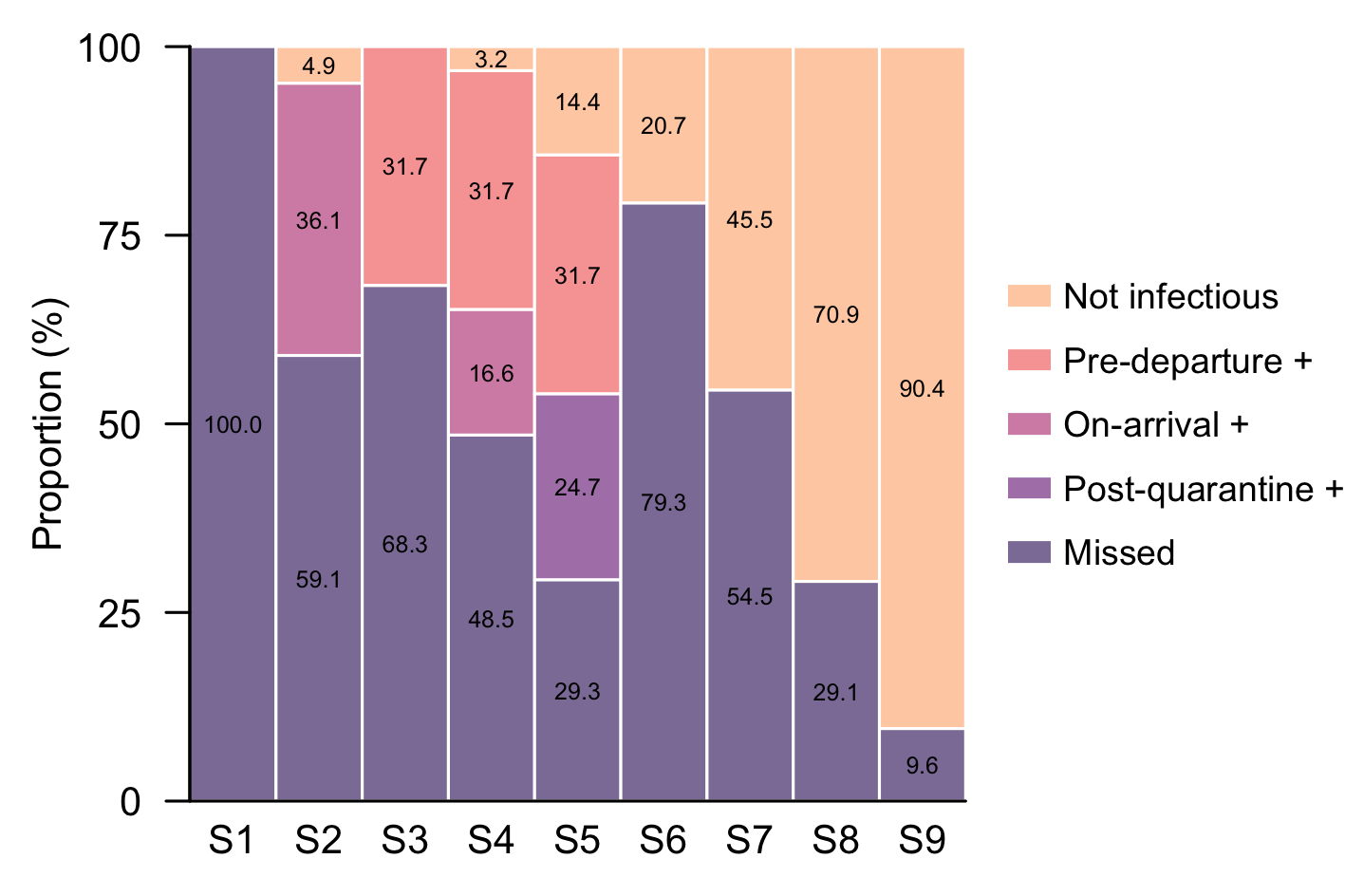

**Figure S3.** Proportion of cases missed, detected by PCR tests, and losing infectiousness during quarantine among infected travellers across Strategy 1–9, when the proportion of infections with skin rashes was 30%. The five sections in the legend bar represent cases who lost infectiousness during quarantine or when awaiting PCR results (‘Not infectious’), cases testing positive before departure (‘Pre-departure +’), upon arrival (‘On-arrival +’), and post-quarantine (‘Post-quarantine +’), as well as cases missed and leaked into the (‘Missed’), respectively.

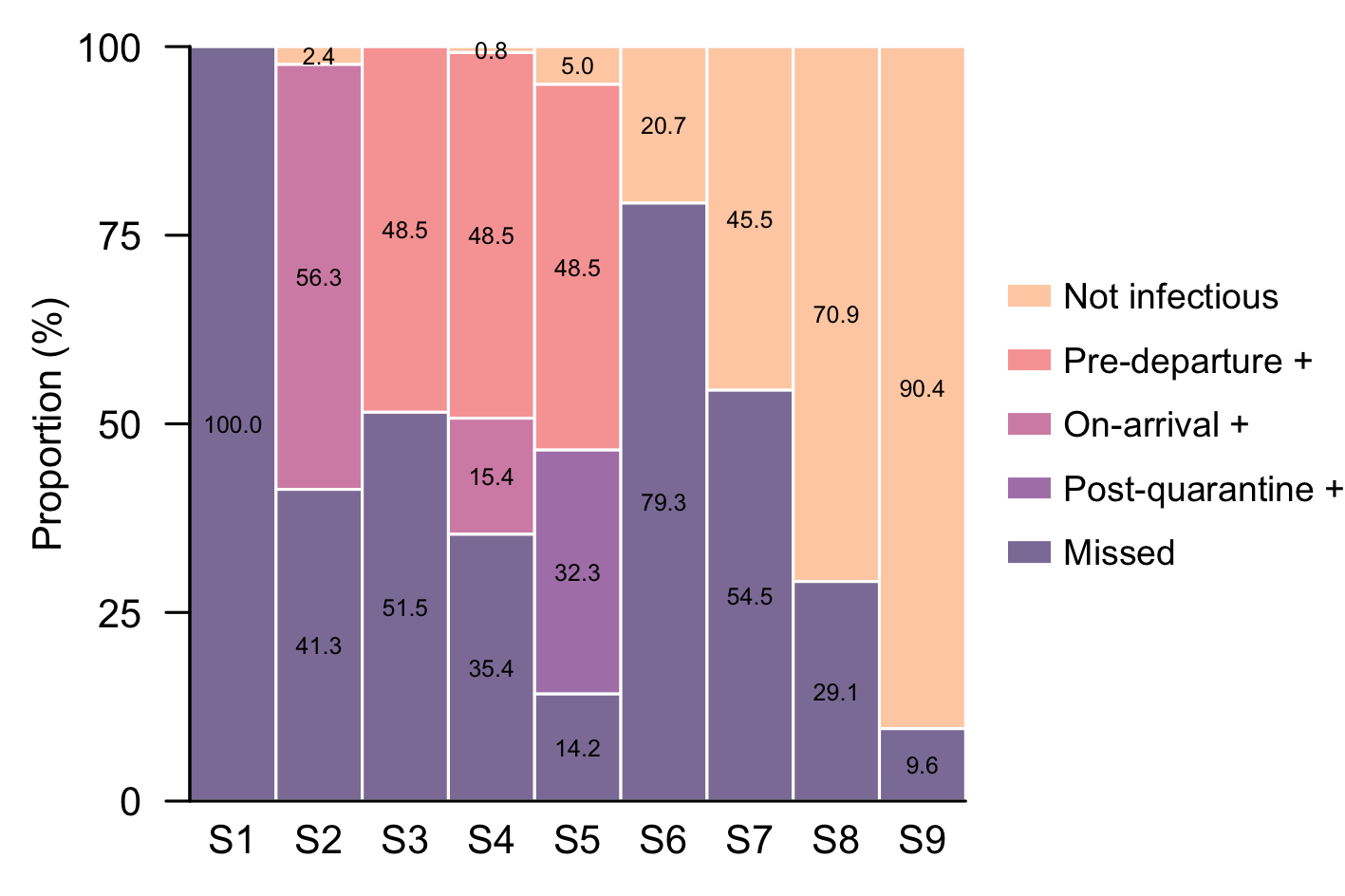

**Figure S4.** Proportion of cases missed, detected by PCR tests, and losing infectiousness during quarantine among infected travellers across Strategy 1–9, when the proportion of infections with skin rashes was 90%. The five sections in the legend bar represent cases who lost infectiousness during quarantine or when awaiting PCR results (‘Not infectious’), cases testing positive before departure (‘Pre-departure +’), upon arrival (‘On-arrival +’), and post-quarantine (‘Post-quarantine +’), as well as cases missed and leaked into the (‘Missed’), respectively.
